# Rapid evaluation of COVID-19 vaccine effectiveness against VOC/VOIs by genetic mismatch

**DOI:** 10.1101/2021.04.22.21254079

**Authors:** Lirong Cao, Jingzhi Lou, Hong Zheng, Shi Zhao, Chris Ka Pun Mok, Renee Wan Yi Chan, Marc Ka Chun Chong, Zigui Chen, Eliza Lai Yi Wong, Paul Kay Sheung Chan, Benny Chung-Ying Zee, Eng Kiong Yeoh, Maggie Haitian Wang

## Abstract

Timely evaluation of the protective effects of COVID-19 vaccines is challenging but urgently needed to inform the pandemic control planning. Based on vaccine efficacy/effectiveness (VE) data of 11 vaccine products and 297,055 SARS-CoV-2 sequences collected in 20 regions, we analyzed the relationship between genetic mismatch of circulating viruses against the vaccine strain and VE. Variations from technology platforms are controlled by a mixed-effects model. We found that the genetic mismatch measured on the RBD is highly predictive for vaccine protection and accounted for 72.0% (*p*-value < 0.01) of the VE change. The NTD and S protein also demonstrate significant but weaker per amino acid substitution association with VE (*p*-values < 0.01). The model is applied to predict vaccine protection of existing vaccines against new genetic variants and is validated by independent cohort studies. The estimated VE against the delta variant is 79.3% (95% prediction interval: 67.0 – 92.1) using the mRNA platform, and an independent survey reported a close match of 83.0%; against the beta variant (B.1.351) the predicted VE is 53.8% (95% prediction interval: 39.9 – 67.4) using the viral-vector vaccines, and an observational study reported a close match of 48.0%. Genetic mismatch provides an accurate prediction for vaccine protection and offers a rapid evaluation method against novel variants to facilitate vaccine deployment and public health responses.

## Main

Vaccination is a crucial measure to control the transmission scale and mitigate the impact of COVID-19 infections. To date, 19 vaccines against SARS-CoV-2 are in early use or have been fully approved for application in mass population^1^. However, protective effect of the various vaccine products is under the challenge of new genetic variants. Vaccine efficacy or effectiveness (VE) against COVID-19, which measures the relative reduction of risk a disease outcome in clinical trials or mass population, exhibited a wide range of variation from 10.4% to 97.2%^2-5^.

A number of reasons contribute to the variation in VE that makes it difficult to directly interpret and inform the protective effect of vaccines. The notable factors include the technology platforms, the target population, differences in study protocols, background risk of COVID-19 and time of study. The various vaccine technology strategies generated non-identical immune correlates of protection for SARS-CoV-2 infection^6^. For instance, the LNP-mRNA vaccine (Moderna) induces S-specific IgG, high T_H_1 cell responses and low T_H_2 cell responses^7,8^, while the inactivated virus strategy (Sinovac) generates S, RBD and N-specific IgG, without obvious T cell responses^9,10^. Among all the influencing factors, emerging genetic variants relative to the vaccine strain play a critical role in affecting vaccine effectiveness. Many investigations showed that neutralizing activity in plasma or sera of vaccinated individuals against variants containing E484K and N501Y mutations decreased significantly^11-13^. Viral structure studies demonstrated that these amino acid substitutions on the S protein may alter virus-host cell interactions and reshape antigenic surfaces of the major neutralizing sites, thus leading to immune evasion^14,15^. While the mechanisms of immune escape caused by the new mutations are continuously being elucidated in experimental studies, an integrative framework to quantify the effect of genetic mismatch on VE would be instrumental for efficient evaluation of vaccine protection for any country in real-time. The genetic mismatch from vaccine strains due to evolution of the circulating strains occurred in different time periods and locations could provide a relatively compact approach to account for the spatial-temporal confounding factors for VE and facilitate the interpretation of vaccine protective effect.

In this study, we drew the connection between genetic mismatch of circulating SARS-CoV-2 viruses and reported COVID-19 VE from population studies. Based on previous bioinformatics approach established for the influenza viruses^16,17^, we further advanced the VE estimation framework for COVID-19 by controlling the clustered random variation of technology platforms using a mixed-effects model. Through extensive analysis of worldwide VE studies and genetic sequences, we showed that a significant proportion of the change in VE could be explained by the genetic factor and provided an efficient framework to evaluate vaccine protection.

## Results

### VEs and genetic mismatch distributions by vaccine platforms

VE and genetic mismatch of the four vaccine platforms are compared in **Figure 1**. Within each vaccine platform, the vaccine effectiveness is generally lower compared to the efficacy outcome (**Figure 1a**); while in terms of genetic mismatch (**Figure 1b, Supplementary Figure S3.1**), the vaccine effectiveness cohort encompasses larger genetic mismatch relative to the vaccine efficacy cohorts. This result indicates that genetic mismatch had increased during the mass vaccination phase compared to the earlier clinical trial periods. Across the technology platforms, vaccine protection (efficacy/effectiveness) shows significant difference (ANOVA *p*-value < 0.001, **Figure 1a**). The mRNA vaccines reported the highest mean VE of 89.2% (95% CI: 86.2 – 92.2, N=18), followed by the protein subunit vaccine 77.9% (range: 49.4 – 96.4, N=3), inactivated vaccine 72.3% (95% CI: 64.3 – 80.3, N=8), and viral-vectored vaccines 66.7% (95% CI: 57.5 – 75.6, N=15). Interestingly, the genetic mismatch of these platforms shows a perfect reverse trend, of which the mRNA vaccines cohorts correspond to the smallest mismatch, and the viral-vector the highest. The genetic mismatch summarizes the deviation of genetic variants with respect to the vaccine strains, accounting for time, location and multiple strain co-circulation, for vaccine evaluation at population level using sequencing data.

**Figure 1.**
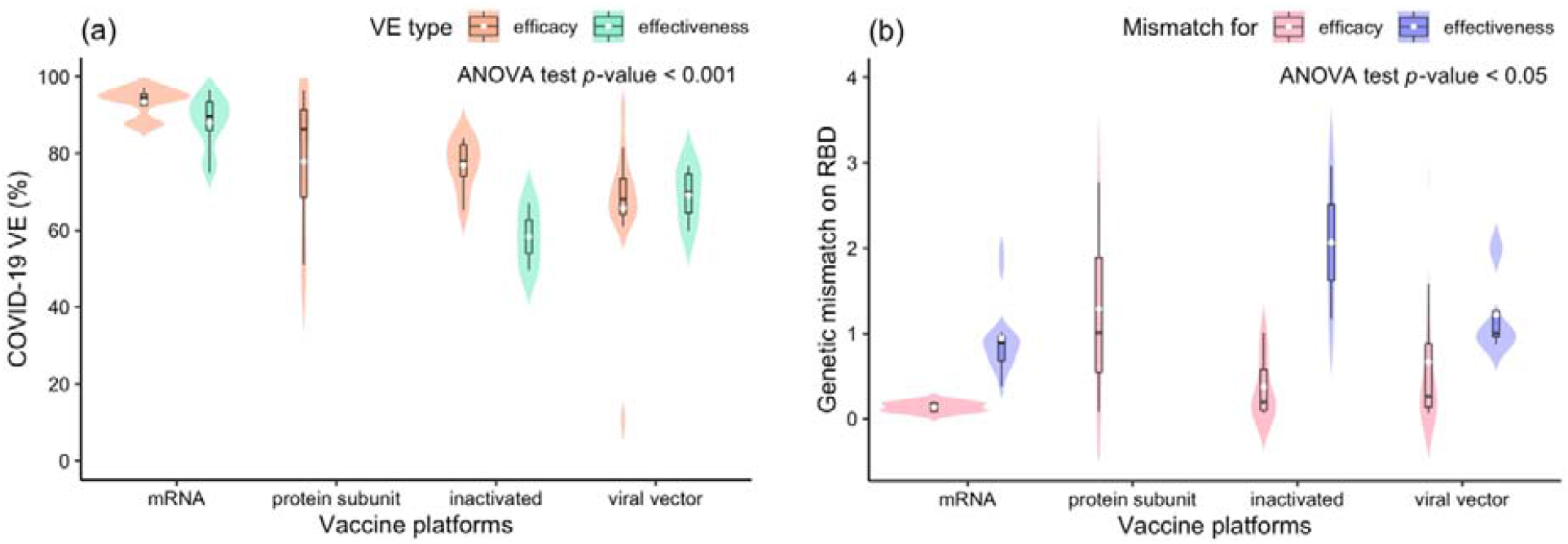
Comparison of COVID-19 vaccine efficacy and effectiveness (VE) and genetic mismatch across vaccine platforms. Panel (a): distribution of the VE estimates for different platforms. The VE of mRNA vaccines is higher than other vaccines (ANOVA *p*-value < 0.001). Panels (b): distribution of genetic mismatch on RBD for different vaccine technologies. Genetic mismatch is the lowest for mRNA vaccines (ANOVA *p*-value < 0.05).

### Relationship between vaccine protection and genetic mismatch

Next, we explored the statistical relationship between vaccine protection and genetic mismatch. Using a mixed-effects model, at most 72.0% of the variations in VE can be explained by the genetic mismatch measure, controlling for the random effect of technology platforms (**Figure 2, Supplementary Table S3.2**). Among the candidate genomic regions, genetic mismatch on the RBD region demonstrated the strongest influence on vaccine protection. For every residue substitution on the RBD, the VE would reduce by an average of 7.2% (95% CI: 3.8 – 10.7, *p*-value < 0.001); the reduction of VE due to one mutation on the NTD and S-protein are 5.4% (95% CI: 2.8 – 7.9) and 1.6% (95% CI: 0.6 – 2.6), respectively (**Supplementary Table S3.2**), while mismatch on ORF1ab, ORF3a, ORF8 and N proteins show no association to VE (**Supplementary Figures S3.3-3.4**). When no genetic mismatch is present, VE is the highest for the mRNA vaccine of an expected level of 94.4% (95% CI: 91.2 – 97.7), estimated by the RBD region; and the inactivated and viral-vector vaccines show a systematically lower VE by 16% and 18.6% relative to the mRNA vaccines.

**Figure 2.**
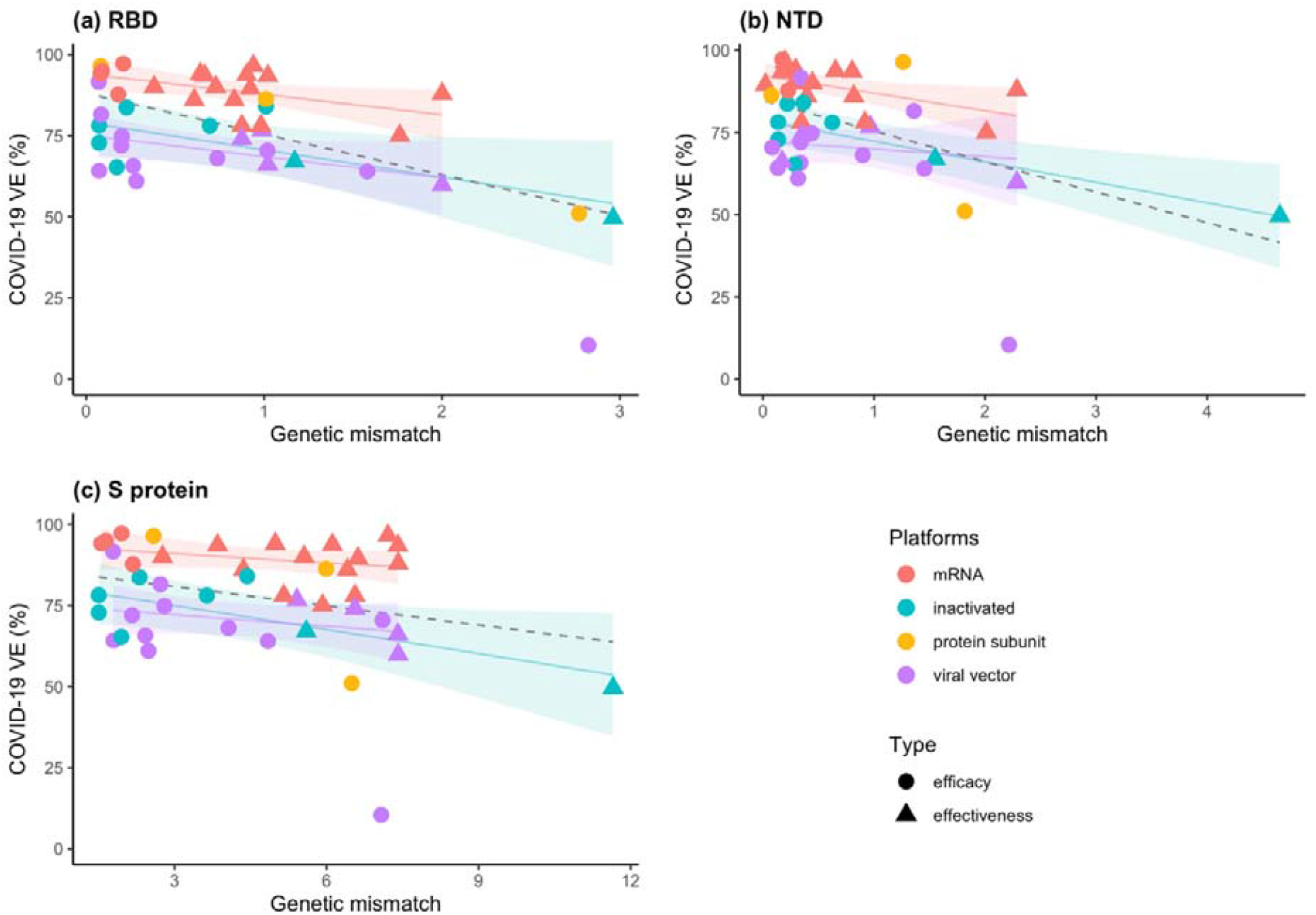
The relationship between VE and genetic mismatch of the circulating SARS-CoV-2 strains to the vaccine strain on S protein. Panels (a-c): negative linear relationships between VE and genetic mismatch for RBD (*p*-value< 0.01, R-sq = 72.0%), NTD (*p*-value <0.001, R-sq = 68.8%), and full-length sequence (*p*-value < 0.01, R-sq= 69.0%), respectively. The dashed line was fitted by all data points. The colored lines were fitted by data points of each platform. The shaded area indicates 95% confidence interval.

### Independent validation and estimating VE against specific genetic variants

In **Figure 3a**, the predicted and observed VEs for all independent datasets are summarized. Calibration plot (**Supplementary Figure S3.5**) shows a close matching, and the concordance correlation coefficient reaches a high level of 0.96 (95% CI: 0.88 – 0.99). These validation results demonstrated feasibility of using genetic mismatch to estimate vaccine performance. In Figure 3b, we further predicted VEs of the mRNA, inactivated and viral-vector vaccines for 15 different variants, including VOC and VOI based on the RBD mismatch (**Supplementary Table S1.3**). Among these variants, four of them have observed VE reported while most of the rest variants have not been surveyed for VE. Against the delta variant (B.1.617.2), the estimated VE is 79.3% (95% prediction interval: 67.0 – 92.1), 63.2% (95% prediction interval: 50.5 – 76.1) and 61.5% (95% prediction interval: 48.3 – 73.4) for the mRNA, inactivated, and viral-vector vaccines, respectively (**Figure 3a**). These estimates are supported by two independent epidemiological studies against the delta variant: the mRNA vaccine BNT162b2 and the viral-vector vaccine ChAdOx1 provided 83% and 61% protection, respectively^18^; and the inactivated vaccine BBV152 conferred 65.2% protection according to^19^. Furthermore, against the beta (B.1.351) and gamma (P.1) variant, the estimated VE for viral-vector vaccines is 53.8% (95% prediction interval: 39.9 – 67.4) and 54.1% (95% prediction interval: 40.0 – 67.7), respectively. An independent study of the viral-vector ChAdOx1-S vaccine reported a VE of 48.0% against these variants^20^.

**Figure 3.**
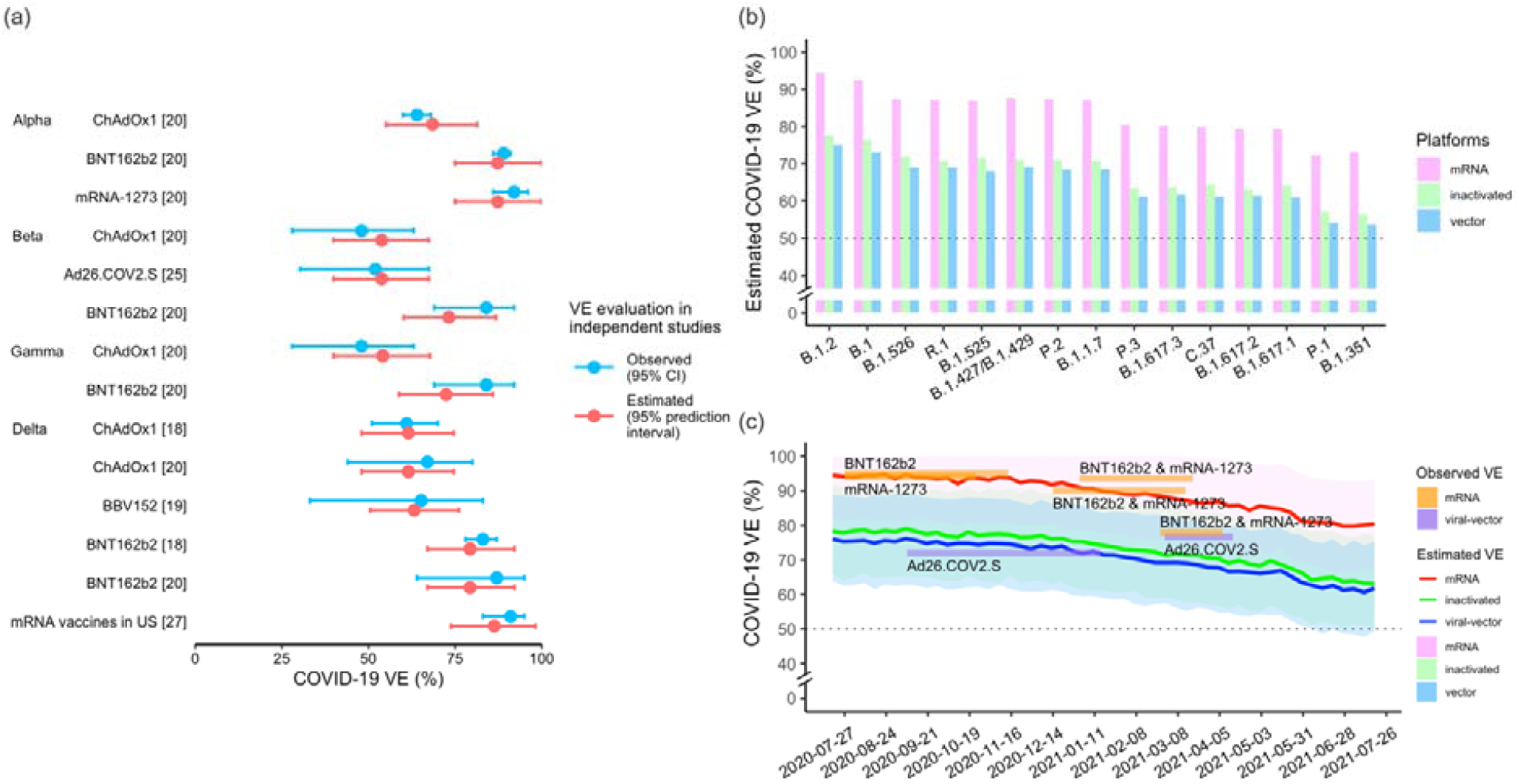
Prediction of the VE based on the genetic distance. Panel (a): Validation outcome of estimated VE and observed VE in independent datasets. The predicted VEs against VOC are close to outcomes of cohort studies observations with concordance correlation coefficient 0.96 (95% CI: 0.88 – 0.99) (Supplementary Figure S3.5). Panel (b): Estimation of the variant-specific VE for mRNA vaccines (pink bar), inactivated vaccines (green bar) and viral-vector vaccines (blue bar). Panel (c): VEs in California were predicted at weekly intervals for different vaccine platforms. The plot indicates that the VE is declining at an accelerating speed. The surveyed VE from clinical trials or observational studies during the same period are overlaid on the trend curve as colored rectangles for reference, and only the mRNA and viral-vector platform vaccines are available. The shaded areas are 95% prediction interval. The dashed line marks the 50% efficacy threshold.

### Depicting trend of VE from the genetic mismatch

VEs are predicted for the major vaccine platforms in California at weekly intervals (**Figure 3c**). In general, an accelerating decreasing trend of VE in California is depicted from the genetic mismatch. We showed that the model can be conveniently applied to track the continuous change of VE. The observed VEs from clinical trials conducted during the period are overlaid on the prediction outcomes for reference^3,21-26^. During February and March 2021, the predicted VE is 86.3% (95% prediction interval: 73.8 – 98.2) for the mRNA vaccines, and an independent survey in the US reported 91% protection for the same vaccine platform^27^.

## Discussion

As novel variants of SARS-CoV-2 keep emerging in the ongoing pandemic, rapid assessment of vaccine performance in populations is crucial to inform public health and clinical responses. This study established an efficient computational framework to estimate COVID-19 VE using virus sequencing data. The predicted VEs against the VOCs are close to outcomes of independent cohort studies. The framework has several advantages. First, it enables prediction of VE against novel variants using existing virus surveillance network to derive a rapid estimate, thus it could inform timely hospital resource allocation and preparedness. Second, it provides an integrated measure to facilitate the interpretation of vaccine effects, which takes account of the confounding effect of time and location related to genetic evolution. Third, through mixed-effects modelling, the framework controls for the random effects in technology platforms, providing a consistent and adaptable prediction framework for inclusion of multiple vaccine platforms.

Among the candidate genomic regions, the RBD and NTD regions exhibit the strongest statistical association with VE. These findings are also supported by biological evidence. The RBD is the major target for neutralizing antibodies that interfere with viral receptor binding^28,29^, and the NTD is reported to be the target of 5-20% of S-specific monoclonal antibodies from memory B cells against SARS-CoV-2^30,31^.

Recent studies have investigated the use of the neutralization titer as a predictor of vaccine efficacy^32^, however the neutralizing results against COVID-19 genetic variants showed varying outcomes. The vaccine protection against the B.1.351 variant reduced from 95.0%^3^ to 75.0%^33^ by BNT162b2. Due to lack of standardized neutralization assays and different protocols, one neutralization study showed that the titer against the B.1.351 variant is 7.6- and 9-fold lower compared to the early Wuhan-related Victoria variant in the BNT162b2 vaccine serum and ChAdOx1 vaccine serum, respectively^12^; while another experiment reported a 2.7-fold decrease in neutralization titers against the B.1.351 strain in the BNT162b2-elicited Serum^34^. The varying neutralization results increase the challenge of inferring vaccine performance solely by neutralization levels. In addition, the association of neutralization with protection across studies showed that neutralizing antibodies might not be deterministic in mediating protection, and the effect of other vaccine-induced immune responses also need to be quantified. This work uses an alternative angle to bridge the link between molecular activities and population level vaccine responses. Further investigations are needed to integrate potential correlates of vaccine protection and improve the existing framework.

The global pandemic of COVID-19 and virus evolution have caused regions in the world to encompass diversified virus populations. We explored the possibility of developing region-specific vaccines and how well they would match the circulating virus profiles. We investigated optimal candidate vaccine strains for 13 regions, including the United Kingdom (UK), Germany, South Africa, Russia, India, Hong Kong (HK), Malaysia, Japan, California, New York, Mexico, Peru and Brazil. Based on the genetic mismatch between vaccine strains and observed viruses circulating in the region and period, hierarchical clustering of the regions was performed to show the similarity of vaccine mismatches (**Figure 4**). We found that no single strain can match to the epidemic viruses in all regions during March-April or May-Jun 2021. Particularly, for the new Moderna vaccine mRNA-1273.351 adopting the B.1.351 variant^35^, the mean genetic discrepancy to local circulating strains is wider compared to either the Wuhan strain or the dominating region strains. This result suggests that updating the vaccine compositions with a single genetic variant might not be sufficient. As manufacturing of region-specific vaccines may not economically feasible, a reconciling strategy might be to provide optimal vaccine candidates for country-clusters that share similar compositions of circulating viruses, or to provide multivalent vaccines.

**Figure 4.**
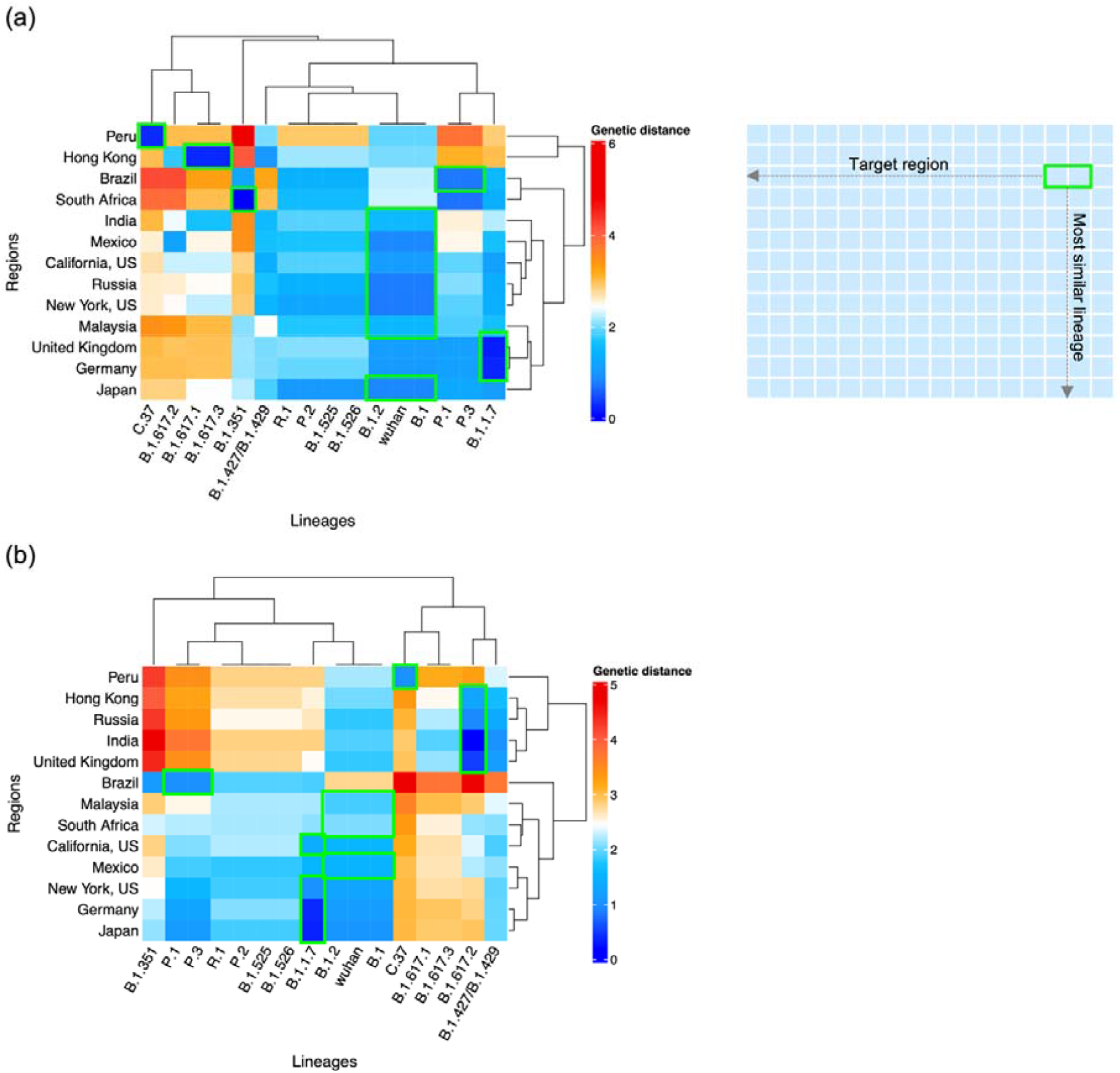
Clustering of regions by circulating strains similarities to VOC/VOIs. Panel (a): Genetic mismatch of genetic variants to the local circulating virus during March and April 2021. The best candidate vaccine antigen for a region measured by genetic distance is highlighted by a green box. Rows: target regions; Columns: candidate vaccine antigens (VOC/VOIs). Panel (b): Genetic mismatch during May and June 2021. For example, in Panel (b), the dark blue of B.1.1.7 in Japan means that the average genetic mismatch between the circulating viruses to the B.1.1.7 is lowest compared to using other variants as vaccine strains, suggesting that the B.1.1.7 is the most optimal vaccine antigen in Japan during May-Jun 2021. The figure shows that no single strain can match to the epidemic viruses in all regions, and the solution might be to provide optimal vaccine candidates for country-clusters that share similar compositions of circulating viruses, or to develop multivalent vaccines.

This study has several limitations. Although the current model reached good statistical significance, the complexity of the model is restricted by the sample size of the available VE studies. Thus, population characteristics and study design factors that may influence VE cannot be included. Secondly, the waned immunity in host was not accounted for in the current model. Thus, the current estimate only suggests the mean protection level within weeks since vaccination based on the data used for model training, and should be interpreted with caution of potentially optimistic estimates. Further study will be sought to consider penalization of the VE according to the time elapsed since last vaccination, as more longitudinal data of immune correlates are available. Thirdly, bias might occur if sequences in databases disproportionately represented regions with known circulation of a given variant. Enhanced efforts are needed to ensure better geographical representativeness of available SARS-CoV-2 sequences. Despite these limitations, the relationship of genetic mismatch and VE observed in multiple countries showed robust outcomes and were validated by independent data. The framework could further pool VE outcomes by various manufacturers using one additional layer of structured modelling, when enough data is available in the future.

To conclude, this work developed a modeling framework integrating data from genetics and epidemiological studies for estimating COVID-19 vaccine effectiveness in a given period and region against a specific variant or for a particular cohort. Rapid assessment of VE before exposure to pathogens can be a useful instrument to inform the vaccine development, distribution and public health responses.

## Methods

### Vaccine Efficacy and Effectiveness Data

Vaccine efficacy is the relative proportion of vaccine protection measured in clinical trials, and vaccine effectiveness is the quantity obtained from observational studies. Both quantities are calculated by (1–RR) ×100, where RR is the relative risk of a COVID-19 outcome in the vaccine group compared to the placebo group. VE reports before May 17, 2021 were collected from journal articles, documents of World Health Organization and Food and Drug Administration, and other government reports. Studies that related to human subjects and contained clear investigation time were included. A total number of 44 VEs were obtained for model building, among which 24 reported vaccine efficacy and 20 surveyed vaccine effectiveness. The vaccine efficacy studies contain thirteen Phase III trials, two Phase II trials, and two Phase II/III trials. Inclusion criteria for the vaccine effectiveness studies are: target population is all age group without special conditions; and the primary outcome is symptomatic COVID-19 infections or confirmed infections requiring medical care. We also extracted 14 VEs from subsequent independent research for validation study. The detailed information of VE for model building and validation is available in **Supplementary Table S1.1-1.2**.

### Genetic Sequences

In the first part of analysis, relationship between genetic mismatch and VE was modelled. Human SARS-CoV-2 strains with collection dates ranging from April 23, 2020 to May 16, 2021 were retrieved from the global initiative on sharing all influenza data (GISAID) EpiCoV database^36^. All available sequences that matched to the period and location of the clinical trials or observational studies were downloaded. A total number of 297,055 full-length genome sequences were sampled from 20 geographical regions for model development. For model validation, a total of 331,116 complete SARS-CoV-2 genome sequences were retrieved from the GISAID.

The source of all SARS-CoV-2 sequences involved in this study was acknowledged in the **Supplementary Acknowledgement Table**. Strains with duplicated names were removed. Multiple sequence alignment was performed using MAFFT (version 7). The ‘Wuhan-Hu-1’ genome (GenBank ‘NC_045512.2’, or GISAID ‘EPI_ISL_402125’) was set as the reference sequence. The variants involved in this study were summarized in **Supplementary Tables S1.3-1.4**. Lineage classification for sequences was referenced from the GISAID.

### Statistical Methods

Following the previous framework developed for influenza virus^16^, let *x* = {*xij*} denote the *i*-th sample from the GISAID database collected for a target population, where *i* =1,…, *n, j*=1,…, *J*; and v = {vj} denote the vaccine strain applied in the target population, where index *j* indicates the *j*-th codon position in the sequence. Denote the amino acids in a given genome region as W= {wk}, where *k* is the index for codon positions contained in the segment, *k* = 1, …, *K*, 0 ≤ *K* ≤ *J*. Suppose the Hamming distance is used as a basic measure of dissimilarity between two sequences, the vaccine genetic mismatch statistic (*d*) calculated for the target population is,

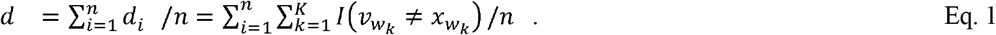

Thus, the *d* summarized the average amino acids mismatch of circulating strains versus the vaccine strain based on a given genome segment in a target population. In this study, we considered a range of candidate *W*, including the receptor-binding domain (RBD), N-terminal domain (NTD), spike (S), nucleocapsid (N), ORF1ab, ORF3 and ORF8 proteins. A schematic representation of SARS-CoV-2 genome and the structure of the S protein are available in **Supplementary Figures S2.1-2.2**. All vaccine strains are based on the Wuhan strain isolated in January 2020. When the target population is composed of subjects infected with multiple co-circulating variants, the *d* captures the viral diversity in the cohort; while when the target population is composed of subjects infects by a specific genetic variant, the mismatch measures variant-specific distance.

In view of the differences in vaccine platforms, a two-level mixed-effects model was adopted to account for the random effect associated with technology platform. We specified the following random-intercept model for a VE outcome (*Y*_*ij*_) of technology *j* and study/trial *i*,

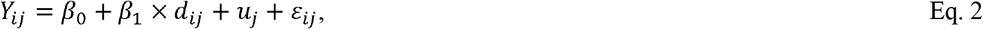

The fixed intercept parameter *β*_*0*_ represents the expected value of VE when genetic mismatch is zero, that is, the maximum protection of a vaccine. *β*_*1*_ represents the fixed effect of genetic mismatch; and *u*_*j*_ denotes the random effect associated with the intercept for platform *j*, which is assumed to follow a normal distribution with zero mean and constant variance 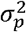. The *εij* denotes the residual of observation from experiment *i* of platform *j*, which follows a normal distribution of zero mean and constant variant *σ*_*2*_. The model was fitted using R package lmerTest^37^. The protein subunit vaccines were excluded in the mixed-effects model as the sample size for this platform is only three. One VE (10.4%) of viral vector vaccine was considered as an outlier and excluded, which is reported from a secondary analysis against the B.1.351 variant in a small-scale South African trial^2^. All analyses were performed using **R** statistical software (version 4.0.3). Statistical significance was declared if *p*-value < 0.05.

Validation study compared the estimated VE of a given platform by using specific lineage sequences or sequences of circulating viruses in the respective regions and periods with fourteen VE outcomes pulling out from independent observational studies. As an application example, we predicted VE against major variants of concern (VOC) and variants of interest (VOI). We also estimated VE of all existing vaccines at weekly intervals from July 20 2020 to July 19 2021 in California to depict the trend of VE through time. The prediction interval of mixed-effects model was calculated using R package merTools^38^.

## Supporting information

Supplementary Acknowledgement Table

## Data Availability

All data used in this work are publicly available.

## Data availability

All data used in this study is publicly available. The detailed information of VE outcomes is available in the supplementary materials. Viral sequence data were downloaded from the global initiative on sharing all influenza data (GISAID) at http://platform.gisaid.org/ and the accession numbers were provided in the supplementary acknowledgment table.

## Code availability

The code is available upon request from the corresponding author.

## Acknowledgements

A complete GISAID acknowledgement table could be found in online supplementary materials. We thank the contributions of all the health care workers and scientists, the GISAID team, and the submitting and the originating laboratories. This work was partially supported by the Health and Medical Research Fund, the Food and Health Bureau, the Government of the Hong Kong Special Administrative Region [COVID190103, INF-CUHK-1], the National Natural Science Foundation of China [31871340, 71974165], and the Chinese University of Hong Kong Grant [PIEF/Ph2/COVID/06, 4054600]. We appreciate the constructive comments from reviewers that improved this work.

## Author Contributions

M.H.W conceived the study, L.C and M.H.W wrote the manuscript. L.C and H.Z. collected data. L.C processed data, carried out the analysis and wrote the first draft. J.L., S.Z., C.K.P.M, R.W.Y.C., M.K.C.C., Z.C., E.L.Y.W., P.K.S.C., B.C.Y.Z. and E.K.Y. critically read and revised the manuscript and gave final approval for publication.

