## Supplementary Materials for "Rapid evaluation of COVID-19 vaccine effectiveness against VOC/VOIs by genetic mismatch"

**S1 Supplementary tables**

### Table S1.1. The source of COVID-19 vaccine efficacy or effectiveness (VE) data by May 17, 2021 for model building

| **No.** | **Developers** | **Vaccine platforms** | **Vaccine products** | **Countries** | **Epidemiological studies** | **VE source** | **Data derived from** |
| --- | --- | --- | --- | --- | --- | --- | --- |
| 1 | Bharat Biotech | Inactivated virus | COVAXIN | India | Phase 3 clinical trial | ^1^ | Text from press release |
| 2 | Sinopharm + China National Biotec Group Co + Beijing Institute of Biological Products | Inactivated virus | Inactivated SARS-CoV-2 vaccine (Vero cell), vaccine name BBIBP-CorV | United Arab Emirates (UAE), Bahrain | Phase 3 clinical trial | ^2^ | Figure 2A |
| 3 | Sinopharm + China National Biotec Group Co + Wuhan Institute of Biological Products | Inactivated virus | Inactivated SARS-CoV-2 vaccine (Vero cell) | UAE, Bahrain | Phase 3 clinical trial | ^2^ | Figure 2A |
| 4 | Sinovac Research and Development Co. Ltd | Inactivated virus | SARS-CoV-2 vaccine (Inactivated) | Brazil | Phase 3 clinical trial | ^3^ | Table 2 |
| 5 |  |  |  | Indonesia | Phase 3 clinical trial | ^4^ | WHO report: Page 7 |
| 6 |  |  |  | Turkey | Phase 3 clinical trial | ^5^ | Text |
| 7 | Novavax | Protein Subunit | SARS-CoV-2 rS/Matrix M1-Adjuvant (Full length recombinant SARS-CoV-2 glycoprotein nanoparticle vaccine adjuvanted with Matrix M) | B.1.351 in South Africa | Phase 2a–b clinical trial | ^6^ | Text |
| 8 |  |  |  | B.1.1.7 in United Kingdom (UK) | Phase 3 clinical trial | ^7^ | Figure 4 |
| 9 |  |  |  | Non-B.1.1.7 in UK | Phase 3 clinical trial | ^7^ | Figure 4 |
| 10 | BioNTech / Pfizer + Fosun Pharma | RNA based vaccine | BNT162 (3 LNP-mRNAs) | Argentina | Phase 3 clinical trial | ^8^ | Table 3 |
| 11 |  |  |  | Brazil | Phase 3 clinical trial | ^8^ | Table 3 |
| 12 |  |  |  | United States of America (US) | Phase 3 clinical trial | ^8^ | Table 3 |
| 13 | Moderna + National Institute of Allergy and Infectious Diseases (NIAID) | RNA based vaccine | mRNA-1273 | US | Phase 3 clinical trial | ^9^ | Figure 3A |
| 14 | Janssen Pharmaceutical | Viral vector (Ad26) | Ad26.COV2.S | Argentina, Chile, Colombia, Mexico, Peru | Phase 3 clinical trial | ^10^ | Table 12 |
| 15 |  |  |  | Brazil | Phase 3 clinical trial | ^11^ | Table 3 |
| 16 |  |  |  | South Africa | Phase 3 clinical trial | ^11^ | Table 3 |
| 17 |  |  |  | US | Phase 3 clinical trial | ^11^ | Table 3 |
| 18 | CanSino Biological Inc./Beijing Institute of Biotechnology | Viral vector (Non-replicating) | Recombinant novel coronavirus vaccine (Adenovirus type 5 vector) | Argentina, Chile, Mexico, Russia | Phase 3 clinical trial | ^12^ | Text from News |
| 19 |  |  |  | Pakistan | Phase 3 clinical trial | ^13^ | Text from News |
| 20 | Gamaleya Research Institute; Health Ministry of the Russian Federation | Viral vector (Non-replicating) | Gam-COVID-Vac Adeno-based (rAd26-S+rAd5-S) | Russia | Phase 3 clinical trial | ^14^ | Table 2 |
| 21 | AstraZeneca + University of Oxford | Viral vector (Non-replicating) | ChAdOx1-S - (AZD1222) | Brazil | Phase 3 clinical trial | ^15^ | Table 2 |
| 22 |  |  |  | B.1.351 in South Africa | Phase 2 | ^16^ | Table 2 |
| 23 |  |  |  | B.1.1.7 in UK | Phase 2/3 clinical trial | ^17^ | Table |
| 24 |  |  |  | Non-B.1.1.7 in UK | Phase 2/3 clinical trial | ^17^ | Table |
| 25 | Sinovac Research and Development Co. Ltd | Inactivated virus | SARS-CoV-2 vaccine (Inactivated) | Brazil | Observational study | ^18^ | Text |
| 26 |  |  |  | Chile | Observational study | ^4^ | WHO report: Page 7 |
| 27 | BioNTech / Pfizer + Fosun Pharma | RNA based vaccine | BNT162 (3 LNP-mRNAs) | England, UK | Observational study | ^19^ | Table 2 |
| 28 |  |  |  | Israel | Observational study | ^20^ | Table 2 |
| 29 |  |  |  | Israel | Observational study | ^21^ | Table 4 |
| 30 |  |  |  | Israel | Observational study | ^22^ | Text |
| 31 |  |  |  | Italy | Observational study | ^23^ | Table 2 |
| 32 |  |  |  | London, UK | Observational study | ^24^ | Text |
| 33 |  |  |  | B.1.1.7 in Qatar | Observational study | ^25^ | Table 1 |
| 34 |  |  |  | B.1.351 in Qatar | Observational study | ^25^ | Table 1 |
| 35 |  |  |  | Sweden | Observational study | ^26^ | Table 2 |
| 36 |  |  |  | B.1.1.7 in UK | Observational study | ^27^ | Table 2-3 |
| 37 |  |  |  | B.1.617.2 in UK | Observational study | ^27^ | Table 2-3 |
| 38 | BioNTech / Pfizer + Fosun Pharma & Moderna + National Institute of Allergy and Infectious Diseases (NIAID) | RNA based vaccine | BNT162 (3 LNP-mRNAs), mRNA-1273 | 33 U.S. states | Observational study | ^28^ | Text from press release |
| 39 |  |  |  | California, US | Observational study | ^29^ | Figure 1 |
| 40 |  |  |  | 8 U.S. locations | Observational study | ^30^ | Table 2 |
| 41 | Janssen Pharmaceutical | Viral vector (Non-replicating) | Ad26.COV2.S | US | Observational study | ^31^ | Table 2 |
| 42 | AstraZeneca + University of Oxford | Viral vector (Non-replicating) | ChAdOx1-S - (AZD1222) | London, UK | Observational study | ^24^ | Text |
| 43 |  |  |  | B.1.1.7 in UK | Observational study | ^27^ | Table 2-3 |
| 44 |  |  |  | B.1.617.2 in UK | Observational study | ^27^ | Table 2-3 |

### Table S1.2. The source of COVID-19 vaccine efficacy or effectiveness (VE) data by August 1, 2021 for validation

| No. | Vaccine products | Vaccine Platforms | Observed VE | (95%CI) | Reference |
| --- | --- | --- | --- | --- | --- |
| Alpha (B.1.1.7) | | | | | |
| 1 | ChAdOx1 | Viral vector | 64 | (60, 68) | ^32^ |
| 2 | BNT162b2 | RNA based vaccine | 89 | (86, 91) | ^32^ |
| 3 | mRNA-1273 | RNA based vaccine | 92 | (86, 96) | ^32^ |
| Beta (B.1.351) | | | | | |
| 4 | ChAdOx1 | Viral vector | 48 | (28, 63) | ^32^ |
| 5 | Ad26.COV2.S | Viral vector | 52 | (30.3, 67.4) | ^11^ |
| 6 | BNT162b2 | RNA based vaccine | 84 | (69, 92) | ^32^ |
| Gamma (P.1) | | | | | |
| 7 | ChAdOx1 | Viral vector | 48 | (28, 63) | ^32^ |
| 8 | BNT162b2 | RNA based vaccine | 84 | (69, 92) | ^32^ |
| Delta (B.1.617.2) | | | | | |
| 9 | ChAdOx1 | Viral vector | 61 | (51, 70) | ^33^ |
| 10 |  |  | 67 | (44, 80) | ^32^ |
| 11 | BBV152 | Inactivated vaccine | 65.2 | (33.1, 83.0) | ^34^ |
| 12 | BNT162b2 | RNA based vaccine | 83 | (78, 87) | ^33^ |
| 13 |  |  | 87 | (64, 95) | ^32^ |
| Independent study on mRNA vaccines | | | | | |
| 14 | BNT162b2 and  mRNA-1273 | RNA based vaccine | 91 | (83,95) | ^35^ |

### Table S1.3. The main circulating variants throughout the world as of July 14, 2021

| **Pango lineages** | **WHO label** | **Nextstrain clade** | **GISAID clade** | **VOI or VOC** | **Earliest documented samples** | **Mutations in S protein** | **Additional amino acid changes monitored** |
| --- | --- | --- | --- | --- | --- | --- | --- |
| B.1 | - | Multiple clades | GK | - | China | D614G | L452R |
| B.1.1.7 | Alpha | 20I/501Y.V1 | GRY | VOC | United Kingdom | 69-70del, 144del, N501Y, A570D, D614G, P681H, T716I, S982A, D1118H | E484K, L452R |
| B.1.2 | - | 20C | GH | - | United States of America | D614G | Q677H |
| B.1.351 | Beta | 20H/501Y.V2 | GH/501Y.V2 | VOC | South Africa | D80A, D215G, Δ242-244, K417N, E484K, N501Y, D614G, A701V | L18F, R246I |
| B.1.427/ B.1.429 | Epsilon | 21C | GH/452R.V1 | Alerts for Further Monitoring | California, United States of America | S13I, W152C, L452R, D614G | - |
| B.1.525 | Eta | 21D | G/484K.V3 | VOI | Multiple countries | Q52R, A67V, 69del, 70del, 144del, E484K, D614G, Q677H, F888L | V144Y |
| B.1.526 | Lota | 21F | GH/253G.V1 | VOI | New York, United States of America | L5F, T95I, D253G, D614G, A701V | D80G, F157S, L452R, S477N, E484K, T859N, D950H, Q957R |
| B.1.617.1 | Kappa | 21B | G/452R.V3 | VOI | India | T95I, G142D, E154K, 157del, 158del, L452R, E484Q, D614G, P681R, Q1071H | V382L, E156G, H1101D, D1153Y |
| B.1.617.2 | Delta | 21A | G/478K.V1 | VOC | India | T19R, 156del, 157del, L452R, T478K, D614G, P681R, D950N | T95I G142D, R158G, A222V, K417N |
| B.1.617.3 | - | 20A | G | - | India | T19R, L452R, E484Q, D614G, P681R, D950N | G142D,E1072K |
| C.37 | Lambda | 21G | GR/452Q.V1 | VOI | Peru | G75V, T76I, 249del, 251del, 252del, L452Q, F490S, D614G, T859N | R203K, D253N |
| P.1 | Gamma | 20J (V3) | GR/501Y.V3 | VOC | Brazil | L18F, T20N, P26S, D138Y, R190S, K417T, E484K, N501Y, D614G, H655Y, T1027I, V1176F | G261S, P681H |
| P.2 | - | 20B/S.484K | GR/484K.V2 | Alerts for Further Monitoring | Brazil | E484K, D614G, V1176F | - |
| P.3 | - | 21E | GR/1092K.V1 | Alerts for Further Monitoring | Philippines | 141-143del, E484K, N501Y, D614G, P681H, E1092K, H1101Y, V1176F | Y265C |
| R.1 | - | 20B | GR | Alerts for Further Monitoring | Multiple countries | W152L, E484K,  D614G, G769V | - |

### Table S1.4. Randomly selected strains from the investigated lineages to plot Figure 4

| **Pango lineages** | **Accession number** | **Strain names** | **Source** |
| --- | --- | --- | --- |
| B.1 | EPI_ISL_1473493 | hCoV-19/England/CAMC-145D216/2021 | GISAID |
| B.1.1.7 | EPI_ISL_1718637 | hCoV-19/England/ALDP-15182AD/2021 | GISAID |
| B.1.2 | EPI_ISL_1553218 | hCoV-19/USA/IL-S21WGS344/2021 | GISAID |
| B.1.351 | EPI_ISL_1534311 | hCoV-19/South_Africa/NHLS-UCT-GS-B188/2021 | GISAID |
| B.1.429 | EPI_ISL_1525760 | hCoV-19/USA/CA-CDC-FG-015454/2021 | GISAID |
| B.1.526 | EPI_ISL_1200537 | hCoV-19/USA/NY-NYCPHL-003648/2021 | GISAID |
| B.1.617.1 | EPI_ISL_1818634 | hCoV-19/India/KA-NIMH-SEQ-374/2021 | GISAID |
| B.1.617.2 | EPI_ISL_1704630 | hCoV-19/India/MH-ICMR-NIV-INSACOG-GSEQ-1304/2021 | GISAID |
| B.1.617.3 | EPI_ISL_1704623 | hCoV-19/India/MH-ICMR-NIV-INSACOG-GSEQ-1294/2021 | GISAID |
| P.1 | EPI_ISL_1495024 | hCoV-19/Brazil/MG-LBI246/2021 | GISAID |
| P.2 | EPI_ISL_1240642 | hCoV-19/Brazil/MG-FUNED-49391-21/2021 | GISAID |
| P.3 | EPI_ISL_1122458 | hCoV-19/Philippines/PH-PGC-02772/2021 | GISAID |
| B.1.525 | EPI_ISL_1729603 | hCoV-19/Germany/BY-RKI-I-095380/2021 | GISAID |
| C.37 | EPI_ISL_1629764 | hCoV-19/Peru/LIM-UPCH-0372/2021 | GISAID |
| R.1 | EPI_ISL_1793651 | hCoV-19/Japan/YCH0165/2021 | GISAID |

**S2 Supplementary figures**

### Figure S2.1. Schematic representation of the SARS-CoV-2 genome

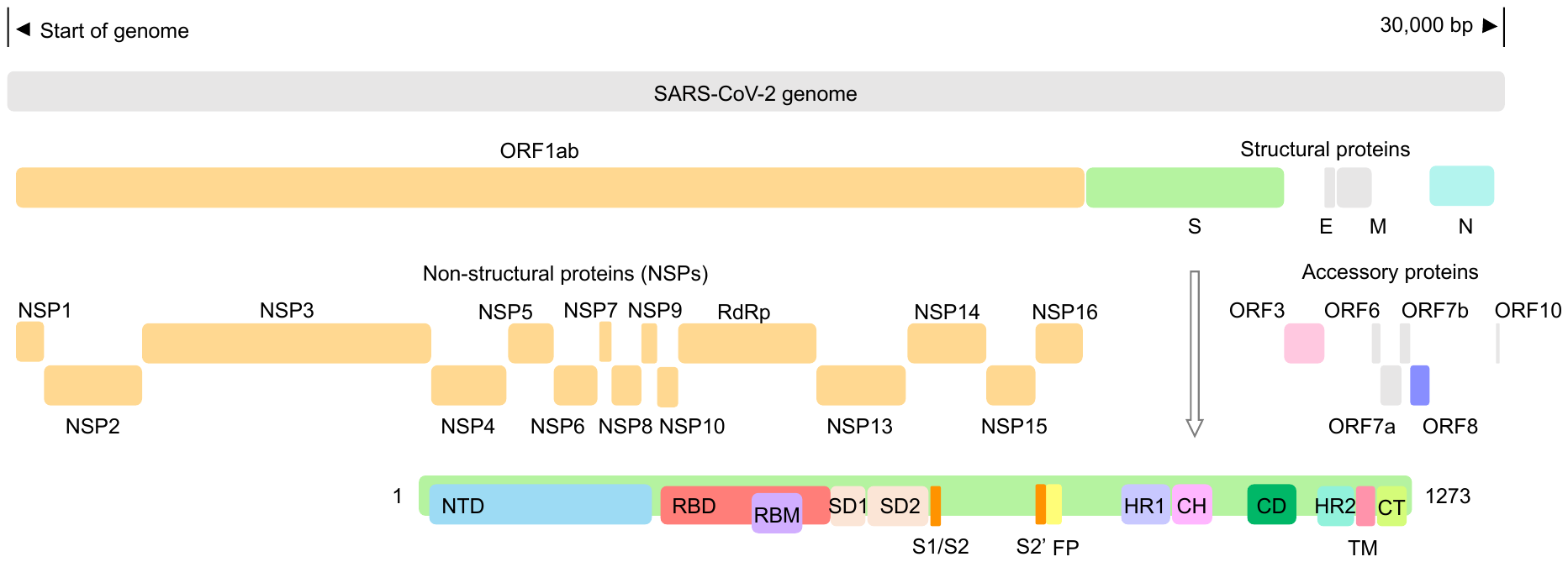

Legend: The proteins highlighted with colours are investigated in this study, including ORF1ab, Spike (S), ORF3a, ORF8 and Nucleocapsid (N) proteins.

### Figure S2.2. Structure of the SAR-CoV-2 Spike protein

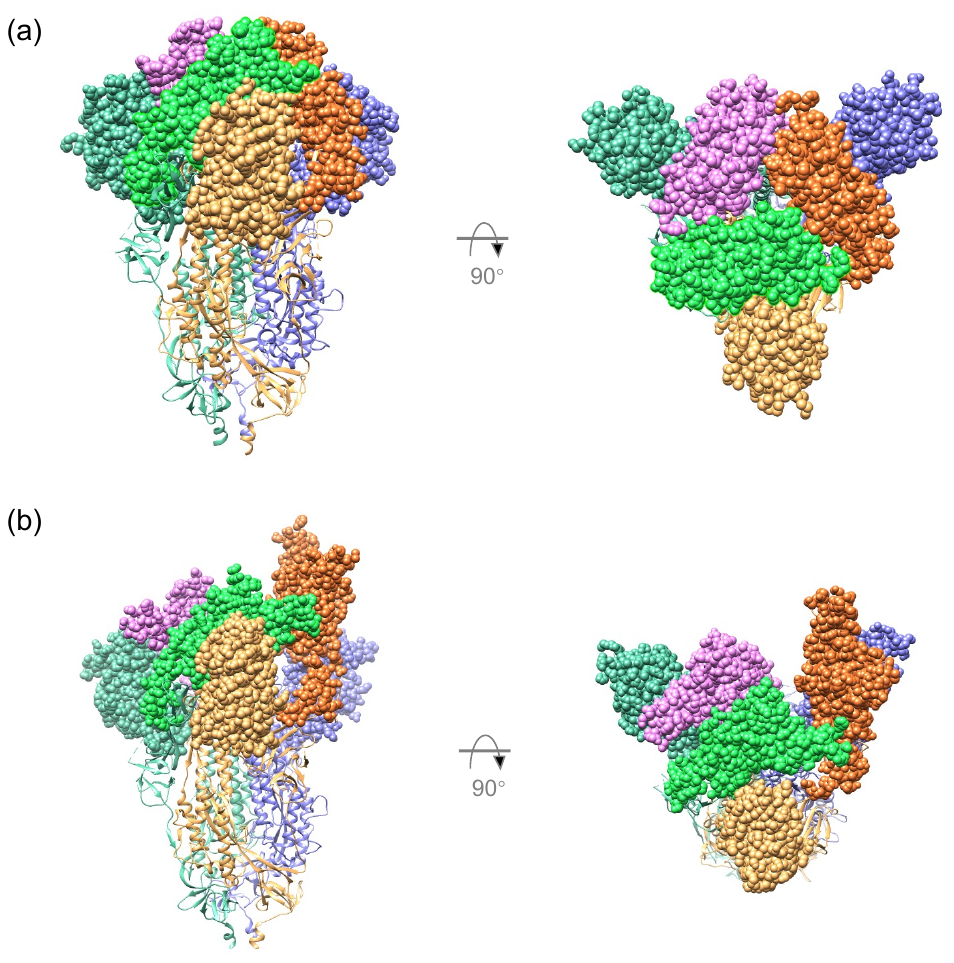

Legend: The location of receptor-binding domain (RBD) and N-terminal domain (NTD) are displayed on the 3D structure of SARS-CoV-2 Spike protein. Panel (a): the conformation of the prefusion trimer; all RBDs in the closed position. Panel (b): the active conformation; one RBD in the open position. The structures are shown from the side view and the top view, respectively. The RBD codons are highlighted in orchid, green and orange spheres. The NTD codons are marked in purple, dark green and yellow. Chimera ^36^ was used to generate the stereo view of S protein structure with 6VXX ^37^ and 7DWZ ^38^ of Protein Data Bank.

**S3 Supplementary results**

### Figure S3.1. Distribution of genetic mismatch on NTD and the complete S protein

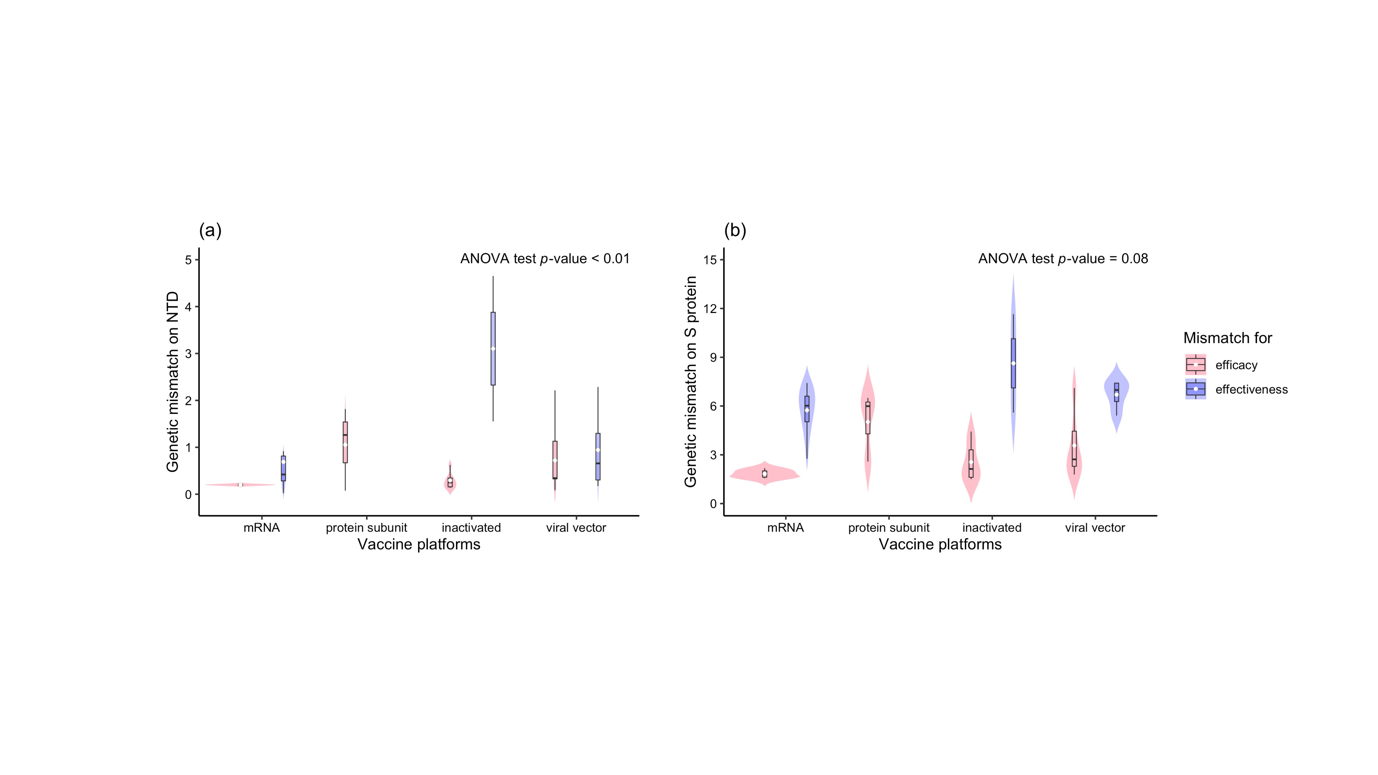

Legend: The genetic mismatch on NTD and S protein was measured. The results show that the genetic mismatch is lowest for mRNA vaccines (ANOVA *p*-value of NTD < 0.05; ANOVA *p*-value of S protein = 0.08).

### Table S3.2. COVID-19 VE prediction models based on the genetic mismatch on RBD, NTD and S protein

| **Genetic regions** | **Vaccine platforms** | **Intercept (95% CI)** | **Slope (95% CI)** | **Random effects variance** | **Error variance** | ***p*-value** | ***R^2^*** |
| --- | --- | --- | --- | --- | --- | --- | --- |
| RBD | mRNA | 94.45 (91.23, 97.66) | -7.24 (-10.70, -3.78) | 106.24 | 49.67 | 0.00222 | 72.00% |
|  | Inactivated | 78.35 (73.6, 83.1) |  |  |  |  |  |
|  | Viral-vector | 75.80 (72.17, 79.43) |  |  |  |  |  |
| NTD | mRNA | 91.97 (88.74, 95.2) | -5.35 (-7.94, -2.76) | 89.11 | 50.44 | 0.00026 | 68.80% |
|  | Inactivated | 77.90 (73.14, 82.66) |  |  |  |  |  |
|  | Viral-vector | 74.61 (70.96, 78.26) |  |  |  |  |  |
| S protein | mRNA | 96.45 (92.98, 99.92) | -1.56 (-2.57, -0.55) | 115.49 | 58.01 | 0.00471 | 69.00% |
|  | Inactivated | 79.00 (73.88, 84.12) |  |  |  |  |  |
|  | Viral-vector | 77.52 (73.6, 81.44) |  |  |  |  |  |

### Figure S3.3. Scatterplot of the observed VE and genetic distance calculated based on different gene segments

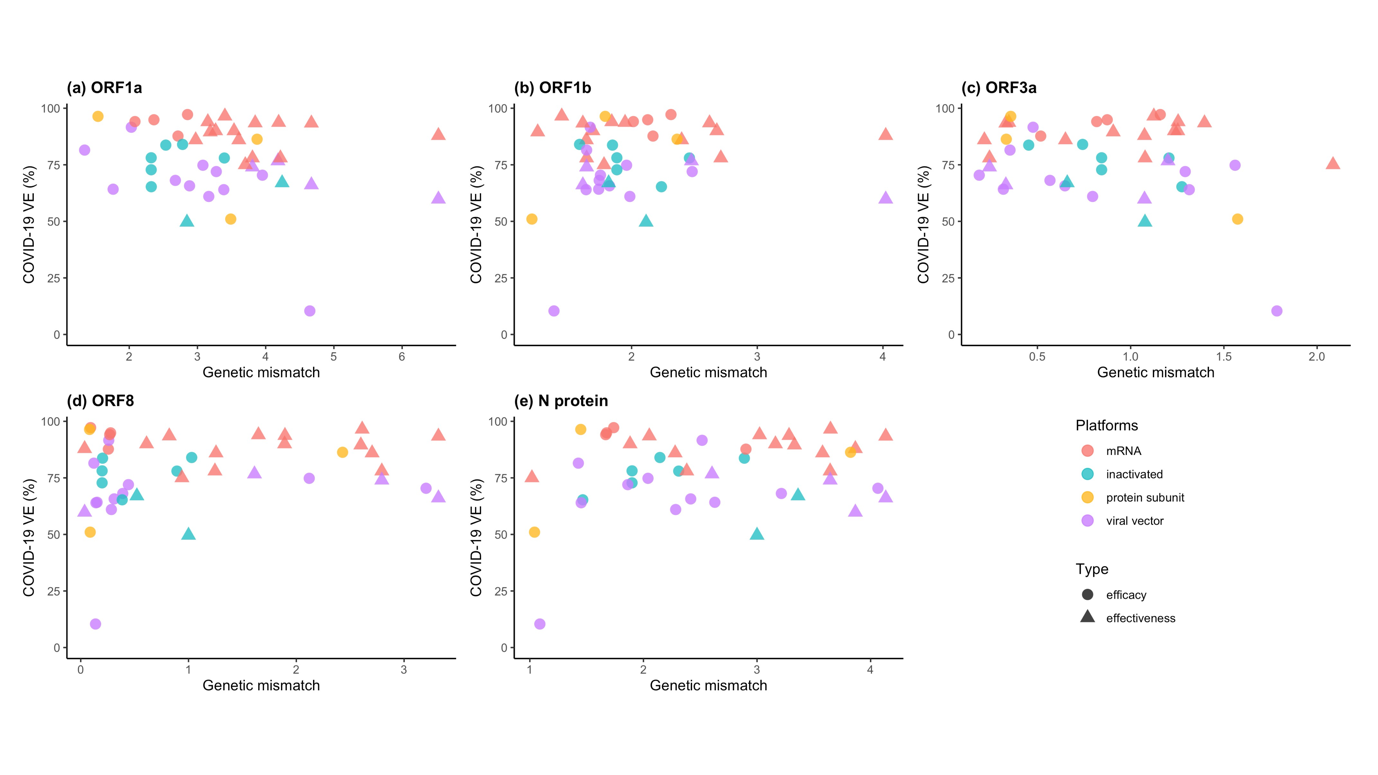

Legend: The same analysis under the null hypothesis to explore the association of VE with genetic distance was performed in ORF1ab, ORF3a, ORF8 and N protein. No significant relationship with VE was observed.

### Figure S3.4. Scatterplot of the observed VE and genetic distance in non-structural proteins (NSPs)

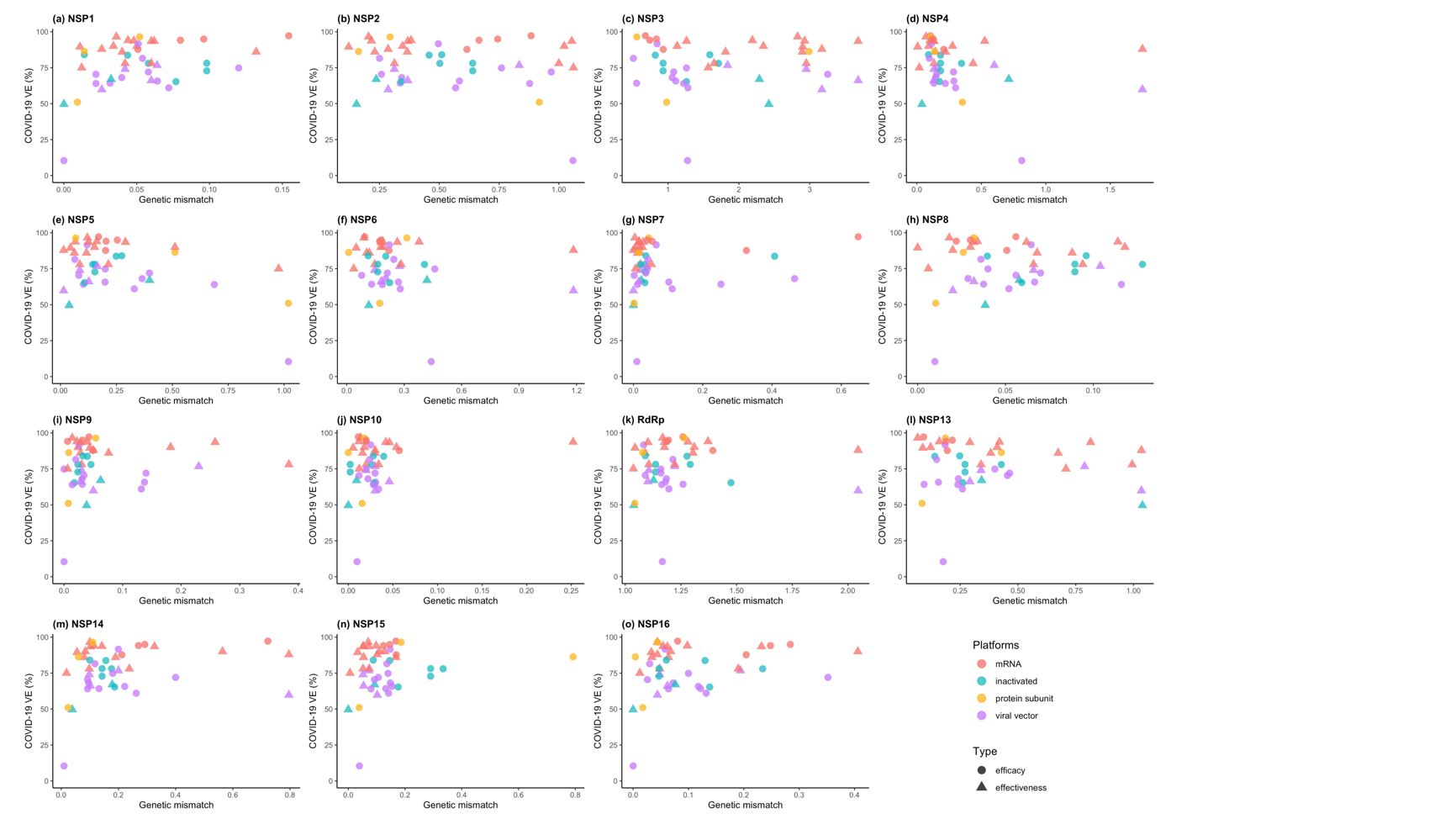

Legend: The ORF1ab polyprotein is composed of 16 non-structural proteins (NSPs). The genetic distance of each NSP was also calculated and no relationships with VE were observed.

### Figure S3.5. Calibration plot for the prediction model of COVID-19 VE in independent studies

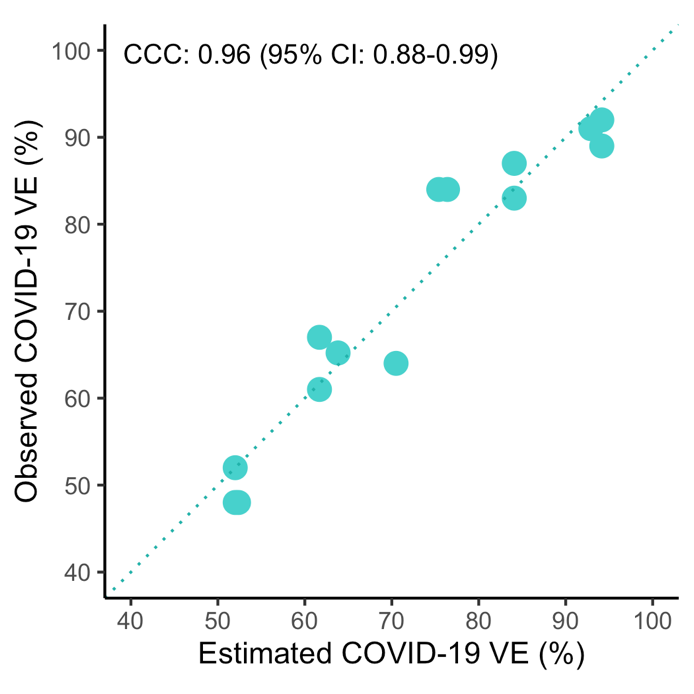

Legend: The observed VEs spread along the 45° line in a long narrow bar and the concordance correlation coefficient (CCC) can reach 0.96 (95% CI: 0.88 – 0.99), suggesting that these VEs are very close to the predicted values.
